# Effectiveness of the ChAdOx1 vaccine in the elderly during SARS-CoV-2 Gamma variant transmission in Brazil

**DOI:** 10.1101/2021.07.19.21260802

**Authors:** Matt D.T. Hitchings, Otavio T. Ranzani, Murilo Dorion, Tatiana Lang D’Agostini, Regiane Cardoso de Paula, Olivia Ferreira Pereira de Paula, Edlaine Faria de Moura Villela, Mario Sergio Scaramuzzini Torres, Silvano Barbosa de Oliveira, Wade Schulz, Maria Almiron, Rodrigo Said, Roberto Dias de Oliveira, Patricia Vieira da Silva, Wildo Navegantes de Araújo, Jean Carlo Gorinchteyn, Jason R. Andrews, Derek A.T. Cummings, Albert I. Ko, Julio Croda

## Abstract

**Background:** A two-dose regimen of ChAdOx1 coronavirus disease 19 (Covid-19) vaccine with an inter-dose interval of three months has been implemented in many countries with restricted vaccine supply. However, there is limited evidence for the effectiveness of ChAdOx1 by dose in elderly populations in countries with high prevalence of the Gamma variant of severe acute respiratory syndrome 2 (SARS-CoV-2).

**Methods:** We conducted a test-negative case-control study to estimate the effectiveness of ChAdOx1 vaccine in adults aged 60 years or older during a Gamma-variant-associated epidemic in São Paulo state, Brazil, between 17 January and 2 July 2021. Cases and matched test-negative controls were individuals, identified from surveillance databases, who experienced an acute respiratory illness and underwent SARS-CoV-2 RT-PCR testing. We used conditional logistic regression to estimate the effectiveness by dose against RT-PCR-confirmed Covid-19, Covid-19 hospitalization, and Covid-19-related death.

**Results:** 61,164 individuals were selected into matched case-control pairs. Starting ≥28 days after the first dose, adjusted effectiveness of a single dose of ChAdOx1 was 33.4% (95% CI, 26.4 to 39.7) against Covid-19, 55.1% (95% CI, 46.6 to 62.2) against hospitalization, and 61.8% (95% CI, 48.9 to 71.4) against death. Starting ≥14 days after the second dose, the adjusted effectiveness of the two-dose schedule was 77.9% (95% CI, 69.2 to 84.2) against Covid-19, 87.6% (95% CI, 78.2 to 92.9) against hospitalization, and 93.6% (95% CI, 81.9 to 97.7) against death.

**Conclusions:** Completion of the ChAdOx1 vaccine schedule afforded significantly increased protection over a single dose against mild and severe Covid-19 outcomes in elderly individuals during widespread Gamma variant transmission.

## Introduction

Multiple vaccines against severe acute respiratory syndrome coronavirus 2 (SARS-CoV-2), the etiologic agent that causes coronavirus disease 19 (Covid-19), have been developed, proven efficacious, and deployed in mass vaccination campaigns^1–3^. Prominent among these vaccines, particularly in lower- and middle-income countries, is the viral vector vaccine, ChAdOx1^4^. Randomized controlled trials (RCT) of ChAdOx1 delivered with a four-week inter-dose interval demonstrated 70.4% (95% CI, 54.8 to 80.6) efficacy against symptomatic Covid-19 in the period starting 14 days after the second vaccine dose^4^, and 64.1% 95% CI, (50.5-73.9) starting at 21 days following the first dose^5^. Based on measured immunogenicity and efficacy following a single dose, many countries have implemented a dose-spacing strategy that uses an inter-dose interval of up to 12 weeks to maximize vaccine coverage^6^ and has been endorsed by the World Health Organization (WHO)^7^.

The emergence of variants of concern (VOC) associated with decreased neutralization activity has created an urgent need to continuously monitor vaccine effectiveness^8^. Recent evidence has suggested reduced effectiveness of a single dose of ChAdOx1 against the Gamma and Delta VOCs^9–11^. Local Gamma VOC transmission has been observed in countries in Latin America which are using ChAdOx1 in mass vaccination^12^. A key question for these countries is the effectiveness of ChAdOx1 by dose against mild and severe Covid-19 outcomes, particularly in priority populations for vaccination such as the elderly.

The Gamma VOC was first detected in the city of Manaus^13^ and has been a driver of Covid-19 resurgence in Brazil and across South America^14^. The Brazilian national immunization program initiated a mass vaccination campaign in January 2021, which administered ChAdOx1 with three-month dose-spacing and provided an opportunity to evaluate vaccine effectiveness following one and two doses during a prolonged Gamma-variant-associated epidemic in São Paulo, the most populous state in Brazil.

## Methods

### Study setting

The study setting and design have been described in detail elsewhere^15,16^. São Paulo State has experienced three Covid-19 epidemic waves, the latest peaking in March 2021, with cumulatively over 3.89 million reported cases, 430,000 hospitalizations, and 130,000 deaths due to Covid-19 as of 9 July 2021^17^ (Figure 1A). During the second and third waves, the Gamma variant increased in prevalence, reaching 80.2% from March to May 2021 among sequenced isolates, to become the predominant circulating variant in the state (Figure 1B). The State Secretary of Health of São Paulo (SES-SP) initiated a mass vaccination campaign on 17 January 2021, prioritizing healthcare workers and elderly populations. Two primary vaccines are being distributed: a two-dose regimen of ChAdOx1, separated by a 12-week interval, and a two-dose regimen of CoronaVac, separated by a two- to four-week interval^18^. As of 9 July 2021, 1.61 million doses of ChAdOx1 (1.11 million first doses and 0.51 million second doses) and 9.07 million doses of CoronaVac (5.62 million first doses and 3.45 million second doses) (Figure 1C) have been administered^19^.

**Figure 1.**
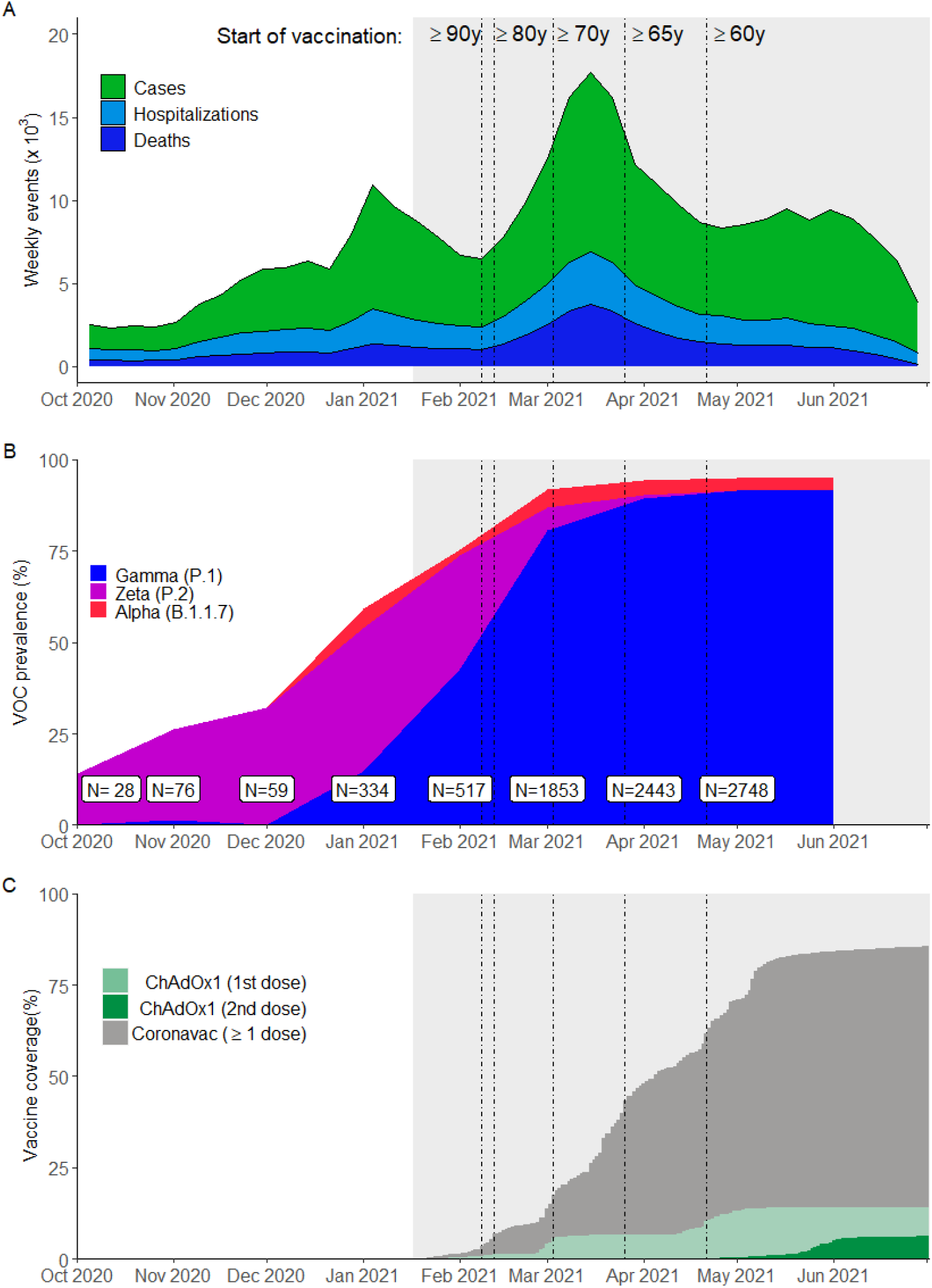
Incidence of reported Covid-19, vaccination coverage, and prevalence of SARS-CoV-2 variants of concern from Oct 1, 2020 to July 2, 2021 in São Paulo State, Brazil. Panel A shows the weekly case count of cases, hospitalizations, and deaths based on positive RT-PCR/Antigen tests for the age group ≥60 years. Panel B shows the monthly prevalence of main SARS-CoV-2 variants of concern among genotyped isolates in the GISAID database^12^ (extraction on July 7 2021). Prevalence was omitted for June and July due to low sample count. Panel C shows daily cumulative vaccination coverage for age group ≥60 years. Population estimates were obtained from national projections for 2020^37^. Vertical lines, from left to right in each panel, show the dates that adults ≥90, 80-89, 70-79, 65-69 and 60-64 years of age in the general population became eligible for vaccination. The gray shaded area represents the study period.

We obtained individual-level information on demographic characteristics, comorbidities, SARS-CoV-2 testing, and Covid-19 vaccination by extracting information on 9 July 2021 from the SES-SP laboratory testing registry, the national surveillance databases for acute respiratory illness (ARI) and severe ARI, and the SES-SP vaccination registry. We retrieved information on SARS-CoV-2 variants from genotyped isolates deposited in the GISAID database^12^. The protocol, including statistical analysis plan, further details of study design, and power calculations, is publicly available (https://github.com/juliocroda/VebraCOVID-19). The study was approved by the Ethical Committee for Research of Federal University of Mato Grosso do Sul (CAAE: 43289221.5.0000.0021).

### Study population and design

The study population was adults ≥60 years of age who had a residential address in São Paulo State and complete and consistent information between data sources on age, sex, residence, and vaccination and testing status and dates. We selected cases who had an ARI, received a positive SARS-CoV-2 RT-PCR test during the study period of 17 January 2021 to 2 July 2021 with sample collection date within 10 days after symptom onset, and without a positive SARS-CoV-2 RT-PCR test in the previous 90 days. We selected test-negative controls who had an ARI, received a negative SARS-CoV-2 RT-PCR test during the study period with sample collection date within 10 days after symptom onset, and without a positive SARS-CoV-2 RT-PCR test in the previous 90 days or following 14 days. Cases and controls who had received a dose of another Covid-19 vaccine before their RT-PCR test were excluded. We matched one control to each case by date of testing (±3 days), age (in 5-year bands), sex, self-reported race (brown, black, yellow, white, or indigenous)^20^, municipality of residence, and prior ARI (defined as at least one previous symptomatic event that was reported to surveillance systems between 1 February 2020 and 16 January 2021).

### Outcomes and Covariates

We estimated the effectiveness of ChAdOx1 against the primary outcome of symptomatic Covid-19 during the period ≥28 days after a single vaccine dose, and 0-13 and ≥14 days after two vaccine doses. Furthermore, we estimated the effectiveness of a single dose during the period 14-27 days after the first dose to understand the onset of protection, and in the period 0-13 days, when the vaccine has no or limited effectiveness^21,22^. An association during this period may serve as a negative control exposure to detect unmeasured confounding in the effectiveness estimate in later time periods^23^. In a secondary analysis, we estimated the vaccine effectiveness following the first dose in the time windows 28-41 days, 42-55 days, and ≥56 days separately. The reference group for vaccination status was individuals who had not received a first vaccine dose before the date of sample collection.

In addition, we estimated vaccine effectiveness against secondary outcomes of Covid-19 hospitalization, ICU admission with Covid-19, mechanical ventilation for Covid-19, and Covid-19-related death. We estimated single-dose effectiveness during the period ≥28 days after the first dose for all outcomes within subgroups defined by age (60-69 years vs. ≥70 years), sex, number of chronic comorbidities (none vs. at least one), reported cardiovascular disease, reported diabetes, and region of residence (“Grande São Paulo” health region vs. others).

### Statistical analysis

We performed conditional logistic regression to estimate vaccine effectiveness for each time window following vaccination. Multivariable models were adjusted for the number of reported comorbidities (categorized as none, one-two, and at least three), previous positive SARS-CoV-2 RT-PCR or antigen test, and age as a continuous variable because we used 5-year age bands as a matching factor. For each outcome, we selected matched pairs in which cases had the outcome of interest, and fit the model described above to each subset.

Our protocol specified that we would conduct proposed analyses after achieving ≥80% power to identify a vaccine effectiveness of 40% against symptomatic Covid-19 ≥28 days after a single dose of ChAdOx1, and 80% power to identify 50% effectiveness of two doses ≥14 days after the second dose. The power was estimated by fitting conditional logistic regressions on 1,000 simulated datasets. After extracting the surveillance databases on July 9 2021 and generating matched case-control pairs, we determined that the power of the study was >99.8% for each analysis and performed the pre-specified analyses. All analyses were performed in R, version 4.0.2.

## Results

### Study population

Among 137,744 individuals eligible for selection as a case or control (Figure 2), 61,164 (44.4%) who provided 61,360 RT-PCR test results were selected into 30,680 matched case and control pairs. Table 1 shows the characteristics of eligible individuals and matched cases and controls. Supplementary Tables 1-3 show the distribution of matched pairs according to vaccination status of cases and controls at the time of RT-PCR testing for the analysis of symptomatic Covid-19, hospitalization, and death. Supplementary Figure 1 shows the timing of discordant pair enrollment, while Supplementary Figure 2 shows the distribution of intervals between administration of vaccine doses and RT-PCR testing.

**Figure 2.**
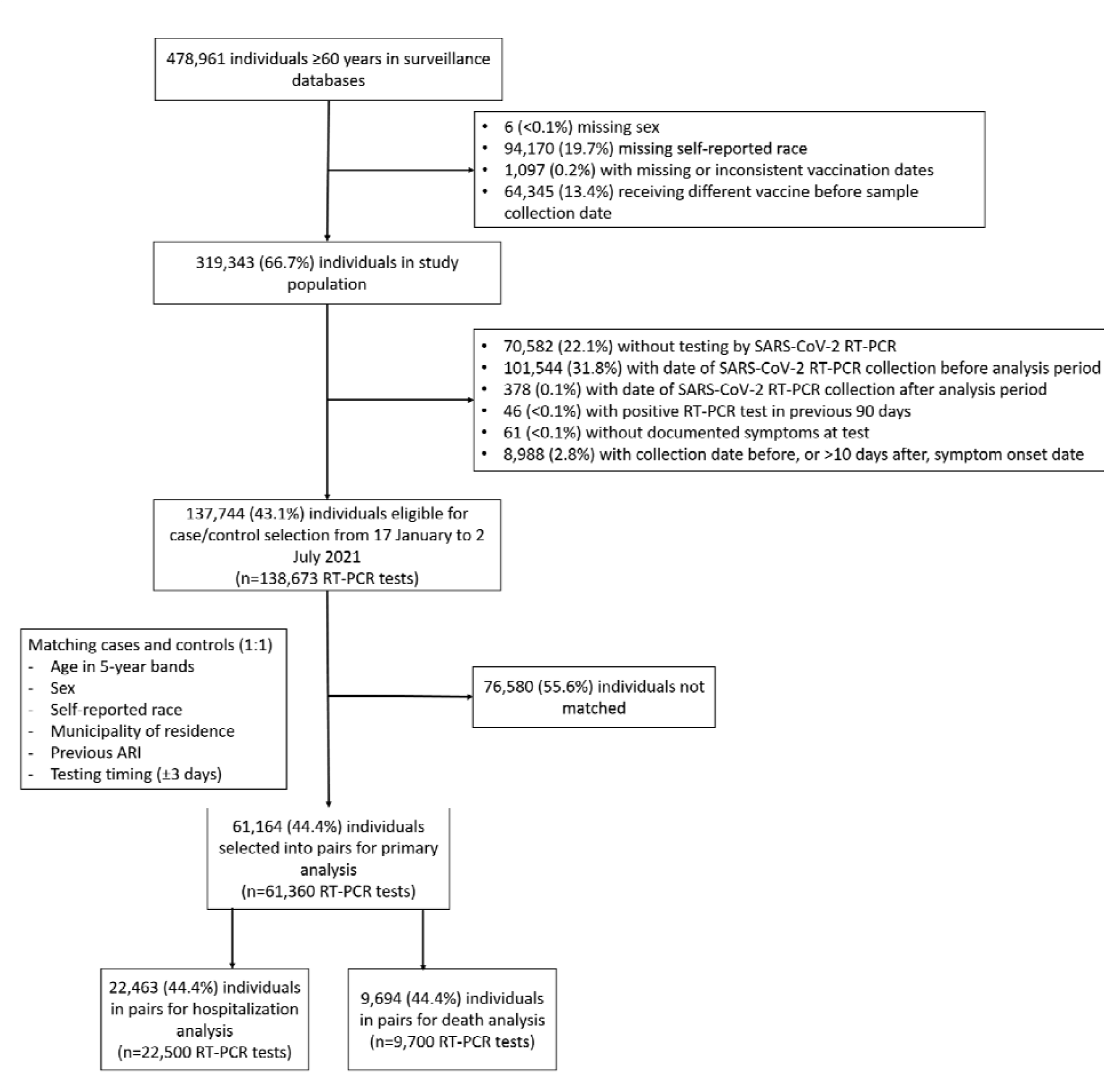
Flowchart of the identification of the study population from surveillance databases and selection of matched cases and controls.

**Table 1.** Characteristics of adults ≥60 years of age who were eligible for matching and selected into case-test negative pairs.

|  | Eligible cases and controls |  | Matched pairs |  |
| --- | --- | --- | --- | --- |
| Characteristics* | Test-negative<br>(n=56,676)<sup>^</sup> | Test-positive<br>(n=81,997)<sup>^</sup> | Controls<br>(n=30,680)<sup>^</sup> | Cases<br>(n=30,680)<sup>^</sup> |
| **Demographics** |  |  |  |  |
| Age, mean (SD), years | 67.90 (7.8) | 67.59 (7.2) | 66.54 (6.5) | 66.55 (6.5) |
| Age categories, n (%) |  |  |  |  |
| 60-69 years | 39,842 (70.3) | 58,181 (71.0) | 23,684 (77.2) | 23,684 (77.2) |
| 70-79 years | 10,346 (18.3) | 15,963 (19.5) | 4,858 (15.8) | 4,858 (15.8) |
| 80-89 years | 5,570 (9.8) | 6,949 (8.5) | 1,966 (6.4) | 1,966 (6.4) |
| $\geq 90$ years | 918 (1.6) | 904 (1.1) | 172 (0.6) | 172 (0.6) |
| Male sex, n (%) | 24,313 (42.9) | 39,180 (47.8) | 12,976 (42.3) | 12,976 (42.3) |
| Self-reported race<sup>†</sup>, n (%) |  |  |  |  |
| White/Branca | 40,860 (72.1) | 58,565 (71.4) | 23,046 (75.1) | 23,046 (75.1) |
| Brown/Pardo | 12,484 (22.0) | 18,463 (22.5) | 6,572 (21.4) | 6,572 (21.4) |
| Black/Preta | 2,720 (4.8) | 4,063 (5.0) | 943 (3.1) | 943 (3.1) |
| Yellow/ Amarela | 605 (1.1) | 890 (1.1) | 119 (0.4) | 119 (0.4) |
| Indigenous/Indigena | 7 (0.0) | 16 (0.0) | - | - |
| Residence in “Grande São Paulo”<br>Health Region, n (%) | 39,767 (70.2) | 53,540 (65.3) | 17,771 (57.9) | 17,771 (57.9) |
| **Comorbidities** |  |  |  |  |
| Reported number<sup>‡</sup>, n (%) |  |  |  |  |
| None | 37,434 (66.0) | 47,262 (57.6) | 20,604 (67.2) | 17,520 (57.1) |
| One or two | 18,121 (32.0) | 32,093 (39.1) | 9,507 (31.0) | 12,136 (39.6) |
| Three or more | 1,121 (2.0) | 2,642 (3.2) | 569 (1.9) | 1,024 (3.3) |
| Cardiovascular disease, n (%) | 13,069 (23.1) | 24,456 (29.8) | 6,865 (22.4) | 9,429 (30.7) |
| Diabetes, n (%) | 8,078 (14.3) | 16,592 (20.2) | 4,308 (14.0) | 6,319 (20.6) |
| **Prior SARS-CoV-2 infection\*\*** |  |  |  |  |
| At least one previous ARI event**, n (%) | 2,722 (4.8) | 1,381 (1.7) | 299 (1.0) | 299 (1.0) |
| Positive SARS-CoV-2 test result<sup>††</sup>, n (%) | 310 (0.5) | 72 (0.1) | 31 (0.1) | 19 (0.1) |
| **Clinical outcomes** |  |  |  |  |
| ARI-related hospitalization, n/n not missing (%) | 7,531/56,468 (13.3) | 30,189/81,515 (37.0) | 3,818/30,581 (12.5) | 11,250/30,502 (36.9) |
| ARI-related death, n/n not missing (%) | 2,571/55,495 (4.6) | 14,082/78,921 (17.8) | 1,321/30,144 (4.4) | 4,850/29,543 (16.4) |
| Interval between symptom onset and RT-PCR testing, median (IQR), days | 3 (2-5) | 4 (3-6) | 3 (2-5) | 4 (2-6) |
| Interval between symptom onset and hospitalization, median (IQR), days | 3 (1-5) | 7 (4-10) | 2 (1-5) | 7 (4-10) |
| Interval between symptom onset and death, median (IQR), days | 8 (4-15) | 16 (11-24) | 8.5 (4-16) | 17 (11-25) |
| **Vaccination status** |  |  |  |  |
| Not vaccinated, n (%) | 44,285 (78.1) | 65,582 (80.0) | 24,868 (81.1) | 25,215 (82.2) |
| Single dose, within 0-13 days, n (%) | 1,877 (3.3) | 3,535 (4.3) | 1,042 (3.4) | 1,141 (3.7) |
| Single dose, 14-27 days, n (%) | 2,543 (4.5) | 4,406 (5.4) | 1,427 (4.7) | 1,380 (4.5) |
| Single dose, ≥28 days, n (%) | 6,918 (12.2) | 7,704 (9.4) | 3,009 (9.8) | 2,731 (8.9) |
| 2nd dose, within 0-13 days, n (%) | 303 (0.5) | 388 (0.5) | 114 (0.4) | 107 (0.3) |
| 2nd dose, ≥14 days, n (%) | 750 (1.3) | 382 (0.5) | 220 (0.7) | 106 (0.3) |
| Interval between 1st and 2nd dose, median (IQR), days | 85 (84-90) | 86 (84-92) | 85 (84-90) | 86 (84-92) |
| Interval between 1st dose and RT-PCR testing, median (IQR), days | 37 (21-57) | 28 (15-48) | 33 (18-50) | 30 (15-48) |
| Interval between 2nd dose and | 21 (12-33) | 13 (8-24) | 20 (10-34) | 13 (8-24) |
| RT-PCR testing, median (IQR),<br>days |  |  |  |  |

\*Continuous variables are displayed as mean (SD); categorical variables are displayed as n (%).
^These numbers refer to RT-PCR tests and represent 120,483 individuals for the eligible cases and controls and 53,495 individuals in the matched cases and controls.
<sup>†</sup>Race/skin color as defined by the Brazilian national census bureau (Instituto Nacional de Geografia e Estatísticas).<sup>20</sup>
<sup>‡</sup>Comorbidities included: cardiovascular, renal, neurological, hematological, or hepatic comorbidities, diabetes, chronic respiratory disorder, obesity, or immunosuppression.
<sup>\*\*</sup>Prior to the start of the study on 17 January, 2021 and after systematic surveillance was implemented on 1 February, 2020.
<sup>\*\*</sup> Reported illness with Covid-19 associated symptoms in the eSUS and SIVEP-Gripe databases.
<sup>††</sup> Defined as a positive SARS-CoV-2 RT-PCR or antigen detection test result.

### Vaccine effectiveness

The adjusted effectiveness of a single dose of ChAdOx1 against symptomatic Covid-19 was 33.4% (95% CI, 26.4 to 39.7) for the period ≥28 days after administration of the first dose (Table 2). The effectiveness of a single dose reached a plateau after 28 days (Figure 3), with no increase observed in later time periods. The adjusted effectiveness of the full two-dose schedule against symptomatic Covid-19 was 38.1% (95% CI, 11.9 to 56.5) in the period 0-13 days after administration of the second dose, and 77.9% (95% CI, 69.2 to 84.2) in the period ≥14 days after administration of the second dose. The estimated effectiveness in the period 0-13 days following the first dose, which serves as a negative control exposure to indicate bias, was -7.1% (95% CI, -19.6 to 4.1). Increasing number of comorbidities were significantly associated with increased odds of Covid-19 in the adjusted analyses (aOR 1.54, 95% CI, 1.49 to 1.60, for one-two comorbidities, and aOR 2.20, 95% CI, 1.98 to 2.45, for three or more comorbidities compared to no comorbidities). A previous positive SARS-CoV-2 viral test was associated with lower odds of Covid-19 (aOR 0.65, 95% CI, 0.37 to 1.17).

**Table 2:** Adjusted effectiveness of a ChAdOx1 against clinical Covid-19 outcomes in adults ≥60 years of age.

|  | Symptomatic Covid-19<br>(n pairs=30,680) |  | Covid-19 hospitalization<br>(n pairs=11,250) |  | ICU admission<br>(n pairs=4,445) |  | Invasive mechanical<br>ventilation<br>(n pairs=2,672) |  | Covid-19-related death<br>(n pairs=4,850) |  |
| --- | --- | --- | --- | --- | --- | --- | --- | --- | --- | --- |
| Vaccine doses and<br>timing | aVE (95% CI) | p-value | aVE (95% CI) | p-value | aVE (95% CI) | p-value | aVE (95% CI) | p-value | aVE (95% CI) | p-value |
| Single dose, within 0-13<br>days vs. unvaccinated* | -7.1% (-19.6-4.1) | 0.23 | 13.1% (-4.4-27.7) | 0.13 | -9.1% (-46.6-18.8) | 0.56 | 12.7% (-29.2-41) | 0.50 | 16.1% (-11.9-37.2) | 0.23 |
| Single dose, 14-27 days<br>vs. unvaccinated* | 17.8% (8.0-26.5) | 0.001 | 33.6% (19.9-45.0) | <0.001 | 39.6% (15.4-56.8) | 0.003 | 51.8% (26.3-68.4) | 0.001 | 37.5% (15.2-54.0) | 0.003 |
| Single dose, ≥28 days vs.<br>unvaccinated* | 33.4% (26.4-39.7) | <0.001 | 55.1% (46.6-62.2) | <0.001 | 50.9% (33.6-63.8) | <0.001 | 70.5% (54.9-80.8) | <0.001 | 61.8% (48.9-71.4) | <0.001 |
| Two doses, within 0-13<br>days vs. unvaccinated* | 38.1% (11.9-56.5) | 0.01 | 59.2% (32.4-75.4) | 0.001 | 50.9% (-41.8-83) | 0.19 | 75.2% (-18.7-94.8) | 0.08 | 77.8% (49.1-90.3) | <0.001 |
| Two doses, ≥14 days vs.<br>unvaccinated* | 77.9% (69.2-84.2) | <0.001 | 87.6% (78.2-92.9) | <0.001 | 89.9% (70.9-96.5) | <0.001 | 96.5% (81.7-99.3) | <0.001 | 93.6% (81.9-97.7) | <0.001 |

aVE: Adjusted vaccine effectiveness

**Figure 3.**
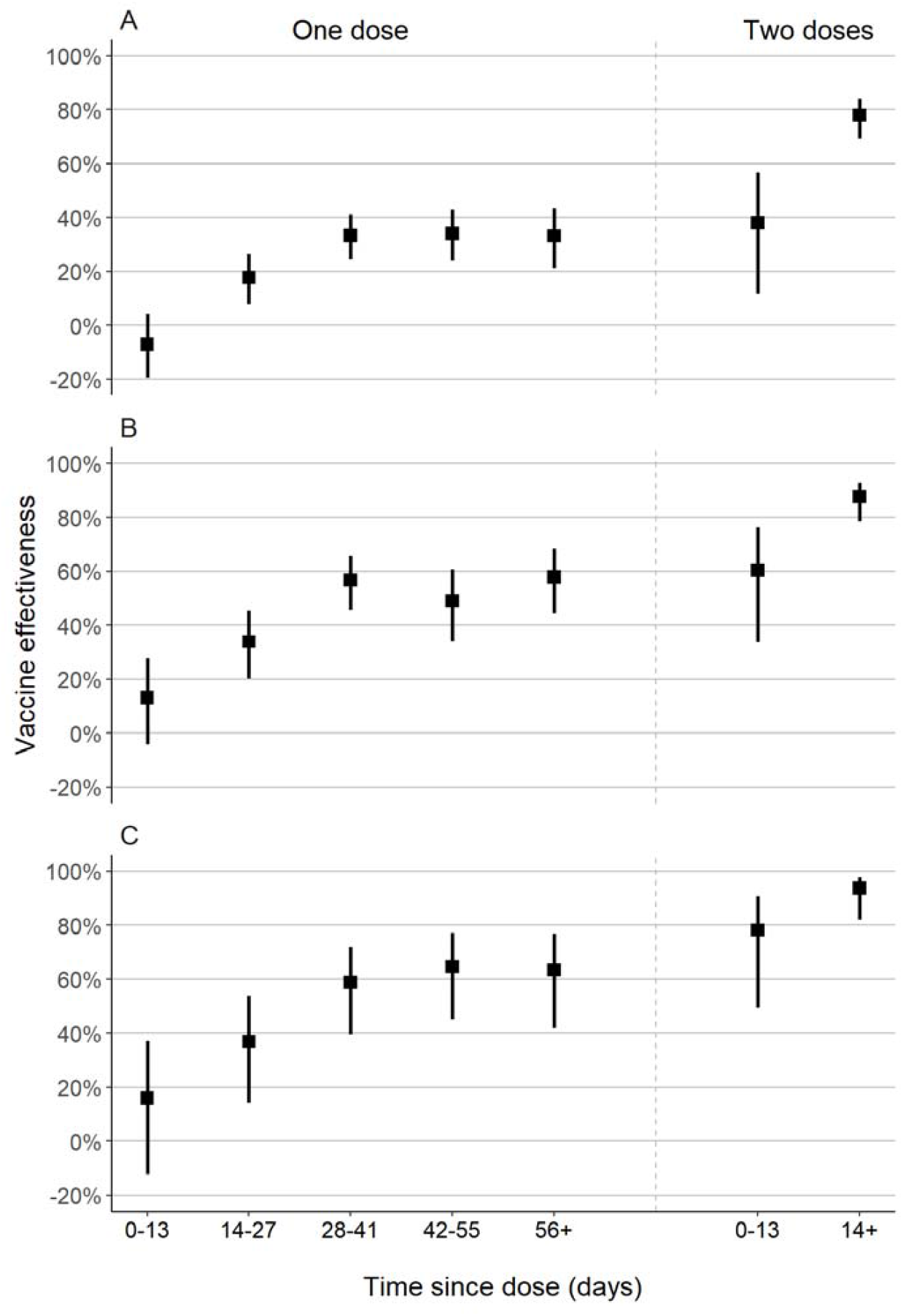
Adjusted vaccine effectiveness of one and two doses of ChAdOx1, by time since vaccination, against symptomatic Covid-19 (A), Covid-19 hospitalization (B), and Covid-19-related death (C)

In the period starting 28 days after the first dose, the adjusted effectiveness of a single dose was 55.1% (95% CI, 46.6 to 62.2) against hospitalization, and 61.8% (95% CI, 48.9 to 71.4) against death (Table 2). The adjusted effectiveness of the two-dose schedule starting 14 days after the second dose was higher: 87.6% (95% CI, 78.2 to 92.9) against hospitalization, and 93.6% (95% CI, 81.9 to 97.7) against death (Table 2). Effectiveness against ICU admission and mechanical ventilation was similar to effectiveness against hospitalization (Table 2). In general, vaccine effectiveness in the “bias-indicator” period 0-13 days after the first dose was low.

The effectiveness of a single dose against symptomatic Covid-19 was lower among those with reported diabetes (24.2%, 95% CI, 11.0 to 35.4) than in those without reported diabetes (35.3%, 95% CI, 28.3 to 41.6) (p_interaction_ = 0.03) (Supplementary Table 4). Similarly, effectiveness was lower among those with at least one reported comorbidity compared to those without a reported comorbidity (Supplementary Table 4). Finally, single-dose effectiveness against hospitalization and death was lower among older individuals, but these analyses lacked sufficient power (Supplementary Table 5).

## Discussion

A key priority for mass vaccination campaigns is to reduce morbidity and mortality in the elderly and other vulnerable populations, especially in the context of limited vaccine supply and VOC emergence. Our test-negative case-control study found that the two-dose schedule of ChAdOx1 in the elderly had robust effectiveness against Covid-19 and severe outcomes during a Gamma-variant-associated Covid-19 epidemic in the period starting 14 days after administration of the second dose: 77.9% (95% CI, 69.2 to 84.2) against symptomatic Covid-19, 87.6% (95% CI, 78.2 to 92.9) against Covid-19 hospitalization, and 93.6% (95% CI, 81.9 to 97.7) against Covid-19-related death. However, a single dose of ChAdOx1 in adults 60 years of age had effectiveness of 33.4% (95% CI, 26.4 to 39.7) against symptomatic Covid-19, 55.1% (95% CI, 46.6 to 62.2) against hospitalization, and 61.8% (95% CI, 48.9 to 71.4) against death. Additionally, no clinically significant effectiveness was detected within 28 days of administration of the first dose.

Randomized controlled trials of ChAdOx1 conducted in multiple countries reported pooled vaccine efficacy of 70.4% (95% CI, 54.8 to 80.6) against symptomatic Covid-19 in the period starting 14 days after the second vaccine dose, and 100% (95% CI, not calculated) against hospitalization for Covid-19^4^. A secondary analysis estimated efficacy of 64.1% (95% CI, 50.5 to 73.9) starting at 21 days following the first dose^5^. Subsequent observational studies have largely supported the effectiveness of ChAdOx1 against symptomatic Covid-19 and hospitalization in elderly populations^21,22,24–26^. In addition, these studies provided further evidence for the effectiveness of a single dose of ChAdOx1 against infection with SARS-CoV-2^26^, symptomatic Covid-19^21^ and hospitalization^22,24,25^, with onset of clinical effectiveness occurring between 21 and 28 days.

Emerging VOCs have been associated with reduced neutralization by serum from individuals who have been infected with non-VOC strains, and vaccinated^27,28^, including those who are elderly^29^, raising the possibility of decreased effectiveness. An RCT of ChAdOx1 conducted in South Africa found no effectiveness, albeit with low precision, of the two-dose vaccine schedule against mild-to-moderate Covid-19 caused by the Beta VOC^30^. Further evidence from observational studies has suggested reduced vaccine effectiveness against symptomatic disease for a single dose of vaccine against Gamma: 48% (95% CI, 28 to 63) after 14 days for ChAdOx1^11^, 61% (95% CI, 45 to 72) after 21 days for mRNA vaccines^31^, and 11% (95% CI, -4 to 23) after 14 days for CoronaVac^16^. However, the complete BNT162b2 schedule has shown robust effectiveness against the Gamma VOC^11^, and a complete schedule of CoronaVac was effective against mild and severe outcomes in settings of widespread Gamma VOC transmission^16^. These findings are consistent with reduced effectiveness of a single dose of BNT162b2 and ChAdOx1 against the Delta VOC observed in the UK^9,10^. Our study adds to this evidence base by estimating single-dose effectiveness of ChAdOx1 over the duration of the inter-dose interval, and demonstrating the substantial benefit of the second dose in elderly individuals in a setting of high Gamma VOC prevalence.

Our findings have implications for vaccination policy in countries experiencing Gamma-variant-associated Covid-19 epidemics. Several countries, including Brazil, are administering the two-dose schedule of ChAdOx1 with a 12-week gap between doses to increase coverage, as WHO currently recommends^7^. The public health benefits of dose-spacing strategies were predicated on robust effectiveness following a single dose^32–34^. In the specific context of VOC emergence and spread, national programs should consider the reduced vaccine effectiveness of a single dose against the Gamma and Delta VOCs in the elderly, together with vaccine supply limitations, speed of vaccination, and logistics, when quantifying the benefits of dose-spacing strategies.

The design of this study lends strength to our findings. The six-month period during which the Gamma variant-associated epidemic and vaccination campaign occurred provided the opportunity to obtain robust estimates of single-dose effectiveness beyond 28 days, and effectiveness of the completed schedule in the same population for direct comparison. The test-negative design reduces bias caused by healthcare-seeking behavior^35^, and we have controlled for additional sources of bias by matching on several predictors of healthcare access and utilization and Covid-19 risk^36^. We used a negative control exposure of vaccine effectiveness within 13 days of receiving the first dose to detect bias in our estimates, and found limited measured effectiveness in this period. This null association suggests that unvaccinated and vaccinated individuals were similar in their underlying risk of Covid-19 and healthcare-seeking behavior^23^. The large sample size allowed us to produce robust estimates even against rare outcomes such as death and to perform subgroup analyses.

Our study has several limitations. We could not estimate the effectiveness against Gamma and non-Gamma Covid-19 cases within this study population, as we did not have access to individual-level genetic data on the virus. In addition, there was likely a proportion of the population that was seropositive without having received a previous positive RT-PCR or rapid antigen test before the study period. These individuals, even if unvaccinated, would be protected from reinfection by natural immunity, thus causing downward bias in our vaccine effectiveness estimates. Controls for the analysis of severe outcomes included controls with mild ARI, who may have had better access to healthcare, leading to bias in our estimates of effectiveness against severe outcomes. Finally, our results cannot be extrapolated to younger populations.

In a setting of widespread transmission of the SARS-CoV-2 Gamma variant, in the general population of elderly individuals, completion of the two-dose schedule of ChAdOx1 afforded a significant increase in protection against mild and severe Covid-19 outcomes compared to a single dose.

## Data Availability

Deidentified databases as well as the R codes will be deposited in the repository https://github.com/juliocroda/VebraCOVID-19

## Author contributions

All authors conceived the study. OTR, MDTH and MD completed analyses with guidance from JRA, DATC, AIK, and JC. MSST, OFPP, OTR and MDTH curated and validated the data. OTR and MDTH wrote the first draft of the manuscript. TLD, RCP, OFPP, EFMV, MA, RS, JCG, WNA provided supervision. All authors contributed to, and approved, the final manuscript. JC is the guarantor. The corresponding author attests that all listed authors meet authorship criteria and that no others meeting the criteria have been omitted.

## Ethics approval

The study was approved by the Ethical Committee for Research of Federal University of Mato Grosso do Sul (CAAE: 43289221.5.0000.0021).

## Acknowledgements

We are grateful for the Pan American Health Organization’s support and the São Paulo State in making the databases available for analysis. JC is supported by the Oswaldo Cruz Foundation (Edital Covid-19 – resposta rápida: 48111668950485). OTR is funded by a Sara Borrell fellowship (CD19/00110) from the Instituto de Salud Carlos III. OTR acknowledges support from the Spanish Ministry of Science and Innovation through the Centro de Excelencia Severo Ochoa 2019-2023 Program and from the Generalitat de Catalunya through the CERCA Program.

## Supplementary appendix

**Supplement to:** Effectiveness of the ChAdOx1 vaccine in the elderly population during a Gamma variant-associated epidemic of Covid-19 in Brazil

**Supplementary Figure 1.**
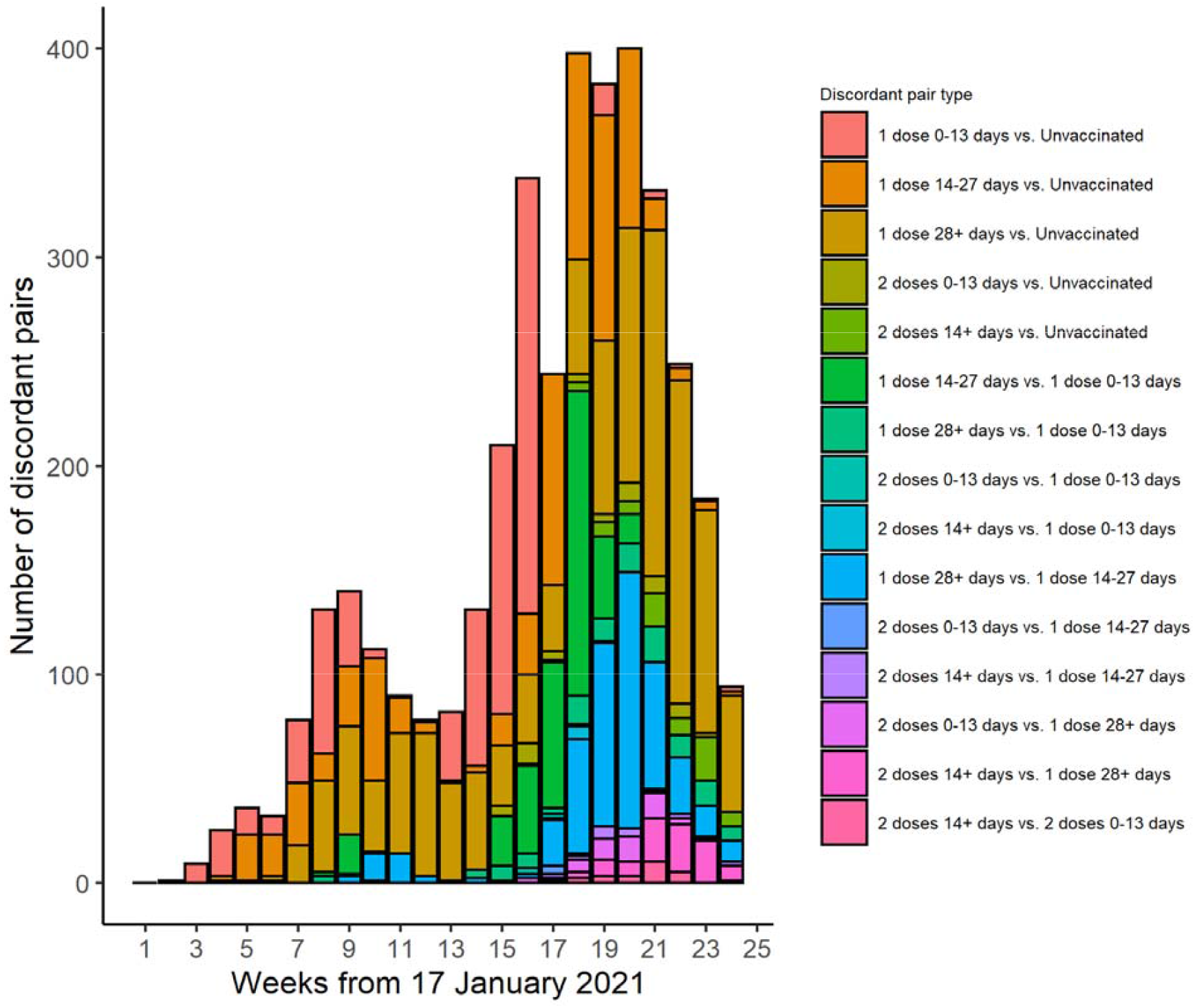
Timing of enrolment of discordant case-control pairs by vaccination category

**Supplementary Figure 2.**
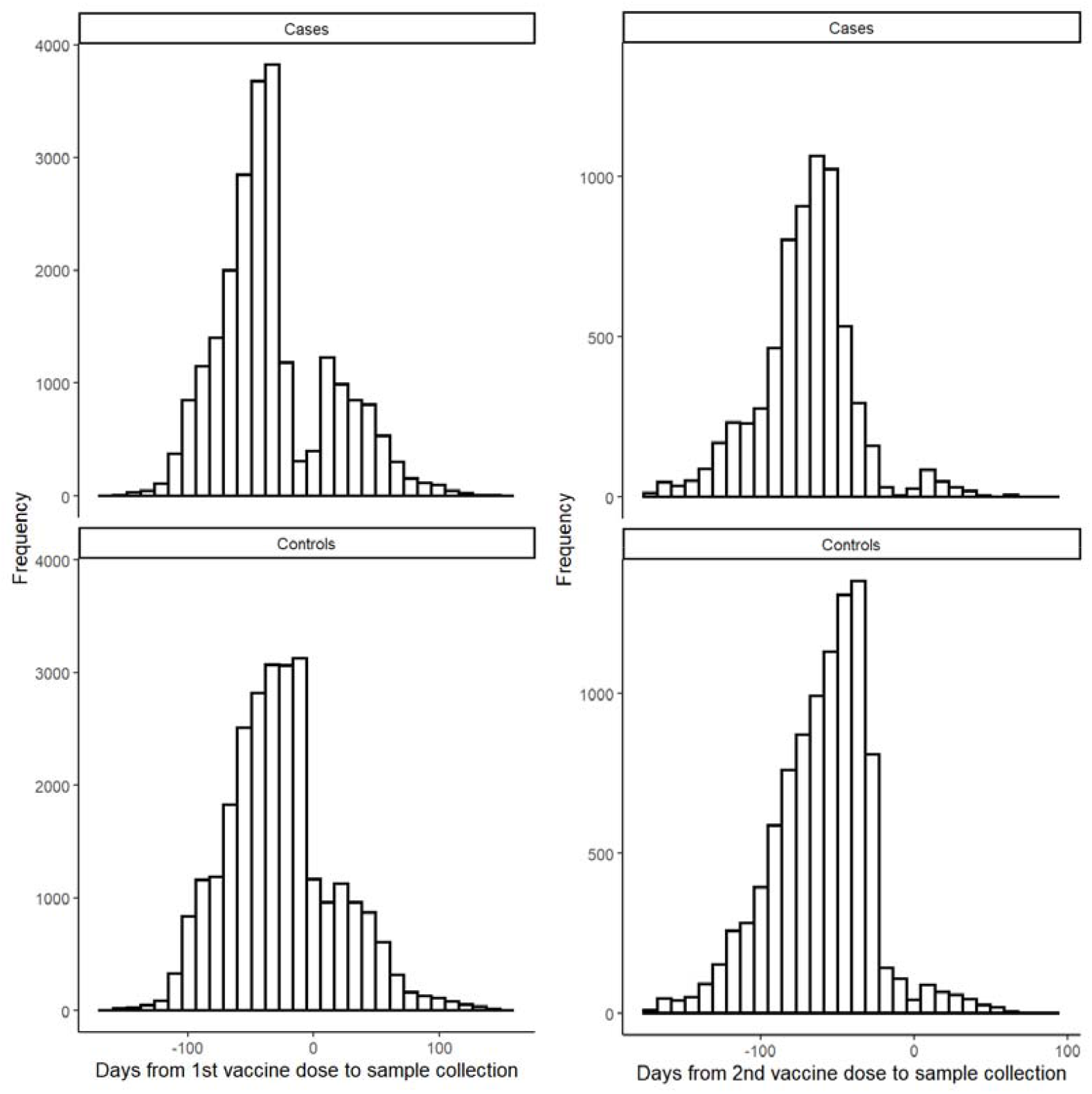
Timing of RT-PCR sample collection date relative to 1^st^ (left column) and 2^nd^ (right column) vaccine dose date, among cases (top row) and controls (bottom row) who were vaccinated during the study period.

**Supplementary Table 1.** Distribution of concordant and discordant matched case-control pairs.

| | Case unvaccinated | Case 1 dose, 0-13 days | Case 1 dose, 14-27 days | Case 1 dose, $\geq 28$ days | Case 2 doses, 0-13 days | Case 2 doses, $\geq 14$ days |
| --- | --- | --- | --- | --- | --- | --- |
| Control unvaccinated | 23,575 | 471 | 313 | 475 | 23 | 11 |
| Control 1 dose, 0-13 days | 463 | 381 | 155 | 41 | 2 | 0 |
| Control 1 dose, 14-27 days | 353 | 205 | 658 | 206 | 1 | 4 |
| Control 1 dose, $\geq 28$ days | 734 | 71 | 232 | 1922 | 25 | 25 |
| Control 2 doses, 0-13 days | 30 | 6 | 6 | 22 | 41 | 9 |
| Control 2 doses, $\geq 14$ days | 60 | 7 | 16 | 65 | 15 | 57 |

**Supplementary Table 2.** Distribution of concordant and discordant matched case-control pairs with hospitalized cases.

| | Case unvaccinated | Case 1 dose, 0-13 days | Case 1 dose, 14-27 days | Case 1 dose, $\geq 28$ days | Case 2 doses, 0-13 days | Case 2 doses, $\geq 14$ days |
| --- | --- | --- | --- | --- | --- | --- |
| Control unvaccinated | 8,454 | 190 | 138 | 163 | 11 | 5 |
| Control 1 dose, 0-13 days | 222 | 143 | 62 | 17 | 0 | 0 |
| Control 1 dose, 14-27 days | 177 | 79 | 221 | 68 | 0 | 0 |
| Control 1 dose, $\geq 28$ days | 369 | 25 | 77 | 632 | 17 | 10 |
| Control 2 doses, 0-13 days | 20 | 4 | 5 | 11 | 21 | 4 |
| Control 2 doses, $\geq 14$ days | 36 | 2 | 8 | 22 | 8 | 29 |

**Supplementary Table 3.** Distribution of concordant and discordant matched case-control pairs with cases who died.

| | Case unvaccinated | Case 1 dose, 0-13 days | Case 1 dose, 14-27 days | Case 1 dose, $\geq 28$ days | Case 2 doses, 0-13 days | Case 2 doses, $\geq 14$ days |
| --- | --- | --- | --- | --- | --- | --- |
| Control unvaccinated | 3,734 | 77 | 54 | 54 | 4 | 2 |
| Control 1 dose, 0-13 days | 103 | 55 | 16 | 7 | 0 | 0 |
| Control 1 dose, 14-27 days | 75 | 30 | 90 | 25 | 0 | 0 |
| Control 1 dose, $\geq 28$ days | 158 | 12 | 30 | 240 | 5 | 2 |
| Control 2 doses, 0-13 days | 9 | 3 | 4 | 6 | 9 | 1 |
| Control 2 doses, $\geq 14$ days | 13 | 2 | 3 | 12 | 4 | 11 |

**Supplementary Table 4.** Adjusted single-dose vaccine effectiveness against symptomatic Covid-19, ≥28 days after first dose, within subgroups

|  | aVE* (95% CI) | p-value for interaction |
| --- | --- | --- |
| **Age** |  |  |
| Age 60-69 years | 34.5% (26.6-41.5) | 0.83 |
| Age $\geq 70$ years | 29.7% (13.3-42.9) | |
| **Sex** |  |  |
| Females | 31.3% (21.6-39.9) | 0.39 |
| Males | 36.0% (25.6-44.9) |  |
| **Comorbidities** |  |  |
| None | 37.0% (29.6-43.5) | 0.02 |
| One or more | 27.0% (17.2-35.6) |  |
| **CVD** |  |  |
| Not reported | 34.9% (27.7-41.5) | 0.16 |
| Reported | 28.7% (18.0-38.0) |  |
| **Diabetes mellitus** |  |  |
| Not reported | 35.3% (28.3-41.6) | 0.03 |
| Reported | 24.2% (11.0-35.4) |  |
| **Health regional area** |  |  |
| “Grande São Paulo” | 28.3% (16.3-38.5) | 0.50 |
| Not “Grande São Paulo” | 37.2% (28.4-44.9) |  |

\* All models adjusted for age as a continuous variable, number of reported comorbidities (none vs. one-two vs. three or more) and previous positive SARS-CoV-2 RT-PCR test

**Supplementary Table 5.** Adjusted one- and two-dose effectiveness against Covid-19 hospitalization and Covid-19-related death by age

|  | Adjusted one-dose<br>VE* (95% CI) | p-value for<br>interaction | Adjusted two-dose<br>VE* (95% CI) | p-value for<br>interaction |
| --- | --- | --- | --- | --- |
| **Covid-19 hospitalization** |  |  |  |  |
| Age 60-69 years | 60.7% (51.1-68.4) | 0.09 | 100.0% (N/A) | <0.001 |
| Age ≥70 years | 41.9% (22.5-56.5) |  | 80.8% (65.2-89.4) |  |
| **Covid-19-related death** |  |  |  |  |
| Age 60-69 years | 72.1% (57.6-81.7) | 0.12 | 100.0% (N/A) | 0.18 |
| Age ≥70 years | 48.0% (22.1-65.3) |  | 89.9% (69.7-96.7) |  |

\* All models adjusted for age as a continuous variable, number of reported comorbidities (none vs. one-two vs. three or more) and previous positive SARS-CoV-2 RT-PCR test

